# Comparative brain metabolomics reveals shared and distinct metabolic alterations in Alzheimer’s disease and progressive supranuclear palsy

**DOI:** 10.1101/2023.07.25.23293055

**Authors:** Richa Batra, Jan Krumsiek, Xue Wang, Mariet Allen, Colette Blach, Gabi Kastenmüller, Matthias Arnold, Nilüfer Ertekin-Taner, Rima F. Kaddurah-Daouk, Alzheimer’s Disease Metabolomics Consortium (ADMC)

## Abstract

Metabolic dysregulation is a hallmark of neurodegenerative diseases, including Alzheimer’s disease (AD) and progressive supranuclear palsy (PSP). While metabolic dysregulation is a common link between these two tauopathies, a comprehensive brain metabolic comparison of the diseases has not yet been performed. We analyzed 342 postmortem brain samples from the Mayo Clinic Brain Bank and examined 658 metabolites in the cerebellar cortex and the temporal cortex between the two tauopathies. Our findings indicate that both diseases display oxidative stress associated with lipid metabolism, mitochondrial dysfunction linked to lysine metabolism, and an indication of tau-induced polyamine stress response. However, specific to AD, we detected glutathione-related neuroinflammation, deregulations of enzymes tied to purines, and cognitive deficits associated with vitamin B. Taken together, our findings underscore vast alterations in the brain’s metabolome, illuminating shared neurodegenerative pathways and disease-specific traits in AD and PSP.

## 1 Introduction

Tau protein hyperphosphorylation and its abnormal accumulation in the brain is a hallmark of several neurodegenerative diseases, including Alzheimer’s disease (AD) and progressive supranuclear palsy (PSP)^1^. The major clinical symptom of AD is dementia, which predominantly impairs memory^2^, while PSP is a Parkinsonian movement disorder^3^. Despite both representing tauopathies, these diseases have several key differences: First, AD is a dual proteinopathy, displaying deposition of both β-amyloid and tau tangles, while PSP is considered to be a pure tauopathy^1^. Second, the specific tau proteins in the two diseases differ in their protein isoforms^4^. While AD presents neuronal tangles composed of tau of both three (3R) and four (4R) microtubule-binding repeat isoforms, shifting from 3R to 4R with disease progression^5^, PSP demonstrates different tau-inclusion patterns predominantly composed of 4R in neurons, astrocytes, oligodendrocytes, and white matter. Further, although both diseases are associated with tau-encoding MAPT haplotypes^6, 7^, distinct genetic risk factors have been discovered for AD^8^ and PSP^7^, signifying divergent genetic components contributing to these two diseases. These differences in the genetic risk, involved proteins, and tau-isoforms might be the basis of the divergent molecular and clinical presentations of these two pathologies.

Comparing the two diseases at the molecular level, we previously reported a high overlap in transcriptomic signatures of these diseases^9, 10^, including dysregulation of biological processes like myelination. As a particularly affected cellular process, metabolic dysregulation involving mitochondrial dysfunction is a common disease pathway in neurodegenerative diseases^11^. Especially in AD pathology, such metabolic modulation is an established key component, affecting cellular processes both in the brain and in the periphery^12–18^. Moreover, our recent study on postmortem human brain tissue suggested tau as the potential driver of AD-associated metabolic alterations in the central nervous system^15^. In the context of PSP, several small-scale studies have indicated the presence of metabolic dysregulation in CSF and in blood^19, 20^.

As AD and PSP share the above-mentioned pathological features, i.e., tauopathy and metabolic alterations, comparing their metabolomic profiles can help identify shared biological pathways that could be targeted for therapeutic interventions. Conducting a thorough brain metabolic comparison of the two diseases is crucial to determine the precise alterations in metabolic homeostasis, which is a hallmark of neurodegenerative diseases^15, 21^. To date, there have been no brain metabolic profiling studies of PSP and no comparative metabolomic studies for these tauopathies.

To address this need, we present a human brain-based metabolic comparison derived from a sizable cohort of neuropathologically diagnosed AD, PSP, and control donors. We analyzed 342 brain samples, comprising 181 from the cerebellar cortex (CER) and 161 from the temporal cortex (TCX). These two brain regions exhibit varying degrees of proteinopathy across the two diseases, with CER being largely unaffected in both^22, 23^ and TCX being affected predominantly in AD^23, 24^. While proteinopathy in the CER of AD patients seems to emerge in later stages^25^, CER has been reported to exhibit disease-driven changes, including atrophy, diffused amyloid presence, and increased microglial activity^26^. From the perspective of PSP, proteinopathy has traditionally only been reported in the anterior CER^27^; however, a recent whole-brain magnetic resonance spectroscopy-based study has revealed metabolic changes across various brain regions including both CER and TCX^28^. Moreover, our previous research has identified transcriptomic alterations in both diseases and brain regions^9, 10^. Hence, we anticipated our analysis will not only identify the metabolomic changes linked with proteinopathies but also the metabolomic dysregulation instigated by neurodegeneration even in the absence of substantial proteinopathy. An overview of the study is depicted in **Figure 1**.

**Figure 1:**
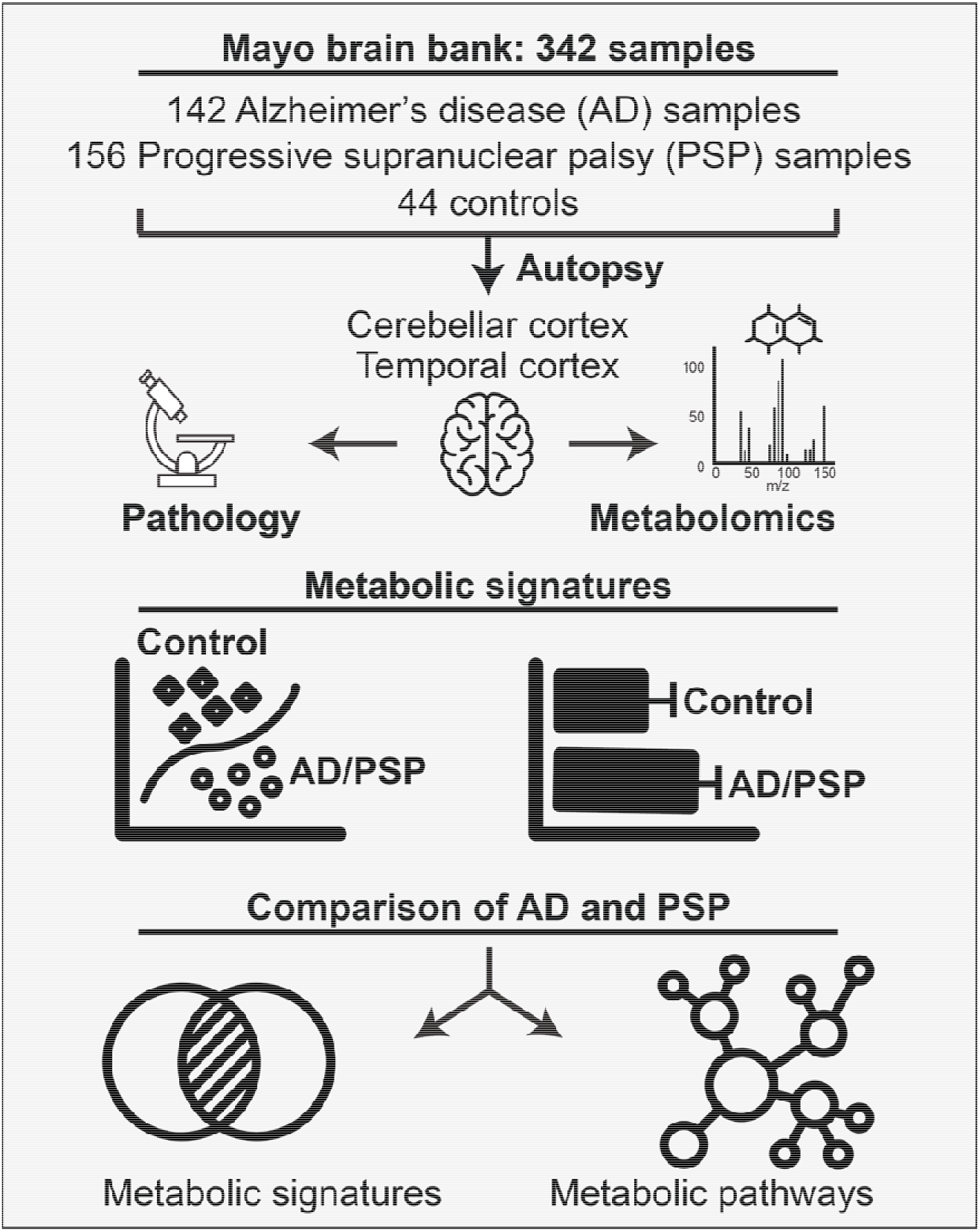
Study overview. 342 Mayo Clinic Brain Bank samples were included in this analysis. For each sample, diagnosis and demographic information were available. Metabolomic profiling was performed on samples from the cerebellar cortex (CER) and temporal cortex (TCX) regions. Metabolic signatures of each diagnosis, i.e., Alzheimer’s disease (AD) and progressive supranuclear palsy (PSP) in comparison to metabolic profiles of controls without neurodegenerative disease. A comparison of the metabolic signatures of the two tauopathies was performed, highlighting the overlap in altered metabolites and metabolic pathways commonly dysregulated in the two diseases.

## 2 Results

### 2.1 Metabolic signatures of the two diseases

We analyzed 342 brain samples from the Mayo Clinic Brain Bank, from Alzheimer’s disease (AD) and progressive supranuclear palsy (PSP) patients as well as controls without neurodegenerative pathologic diagnoses, across two brain regions, cerebellar cortex (CER) and temporal cortex (TCX). Demographic characteristics of the samples across the diagnoses and brain regions are shown in **Table 1**.

**Table 1:** Cohort overview. AD = Alzheimer’s disease. PSP = Progressive supranuclear palsy. IQR = interquartile range, i.e., middle 50% of the value range.

|  | Cerebellar cortex (CER, n = 181) |  |  | Temporal cortex (TCX, n = 161) |  |  |
| --- | --- | --- | --- | --- | --- | --- |
|  | AD<br>(n = 79) | PSP<br>(n = 78) | Control<br>(n = 24) | AD<br>(n = 63) | PSP<br>(n = 78) | Control<br>(n = 20) |
| **Sex** |  |  |  |  |  |  |
| **Males** | 32 | 47 | 11 | 27 | 46 | 8 |
| **Females** | 47 | 31 | 13 | 36 | 32 | 12 |
| **Age at death** | 84<br>(78, 89) | 74<br>(69, 79) | 90<br>(87, 90) | 86<br>(81, 90) | 74<br>(69, 79) | 90<br>(88, 90) |
| **APOE ε4 alleles** |  |  |  |  |  |  |
| **0** | 38 | 66 | 21 | 28 | 67 | 17 |
| **1** | 36 | 11 | 3 | 31 | 10 | 3 |
| **2** | 5 | 1 | 0 | 4 | 1 | 0 |
| <sup>1</sup>n; Median (IQR) |  |  |  |  |  |  |

Samples were profiled using an untargeted LC-MS-based metabolomics platform. This resulted in measurements of 658 metabolites from various metabolic ‘super-pathways’ (**Figure 2a**, **Supplementary Table 1**), covering lipids (44.4%), amino acids (23.3%), nucleotide (7.0%), carbohydrates (5.3%), cofactors and vitamins (3.8%), peptides (2.7%), xenobiotics (2.7%), energy-related metabolites (1.5%), and a series of uncharacterized metabolites (9.3%).

**Figure 2:**
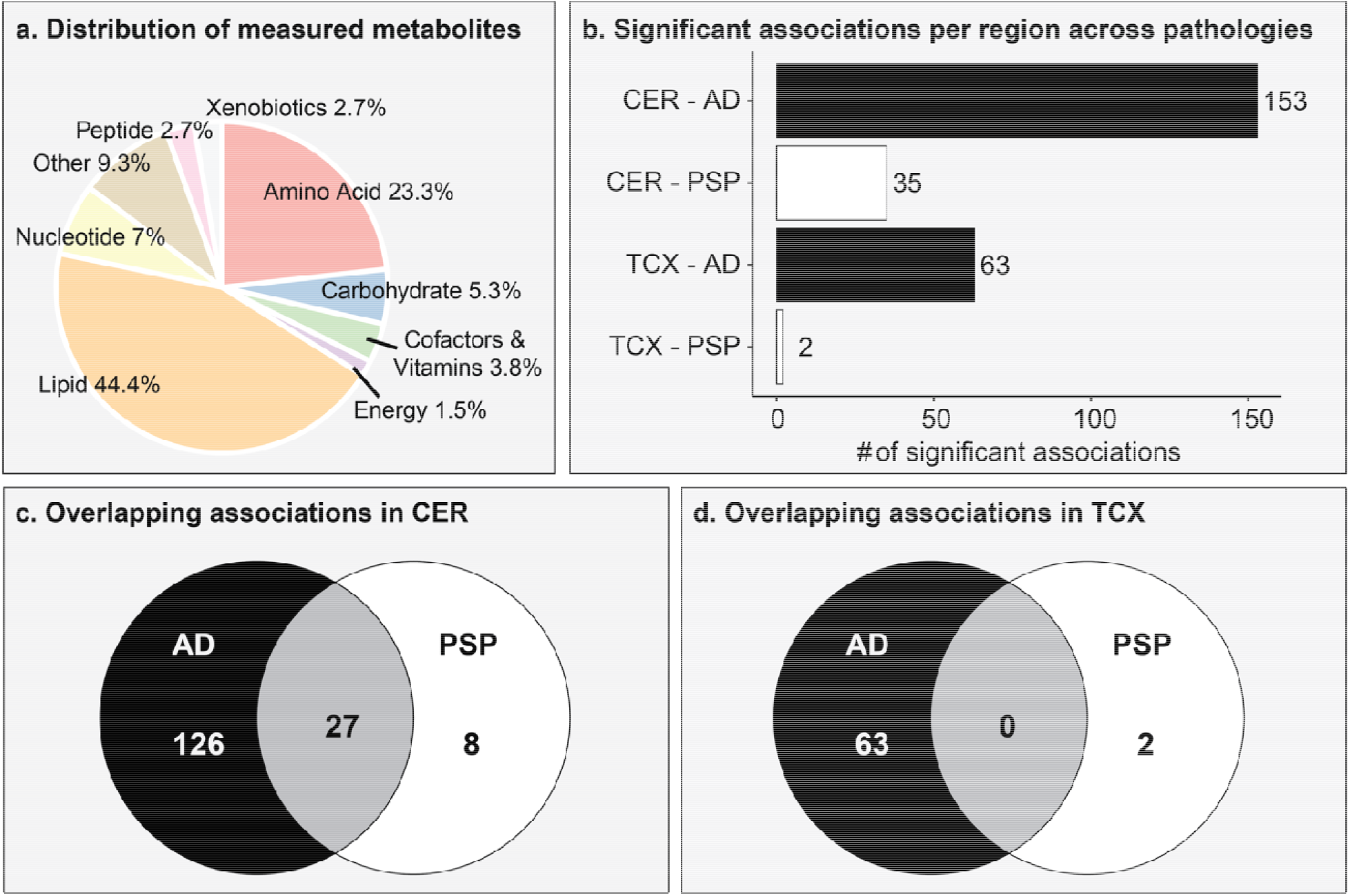
Overview of metabolites and their associations with AD and PSP. a. Distribution of metabolites detected by metabolomics platform across metabolic classes. b. Number of metabolic associations for each brain region and each disease. c. Overlap of metabolic associations between the two diseases in cerebellar cortex samples. d. Overlap of metabolic associations between the two diseases in temporal cortex samples.

To determine the metabolic imprint of the two diseases in the brain, we performed a brain region-wise statistical comparison between disease and control samples. All models were corrected for the number of *APOE* ε4 alleles, age at death, and sex. In the CER region, a total of 153 metabolites were associated with AD, 35 were associated with PSP (**Figure 2b**), of which 27 were commonly associated with both diseases (**Figure 2c**). In the TCX region, 63 metabolites were associated with AD, only 2 were associated with PSP (**Figure 2b**) and none were shared between the two diseases (**Figure 2d**). Detailed statistical results from the two brain regions can be found in **Supplementary Tables 2-5**, respectively.

Overall, AD-related metabolic alterations were observed in both brain regions, while PSP-related alterations had a regional preference for CER and were less pronounced compared to AD-related alterations. Of note, the sample sizes were comparable for both brain regions as well as diseases (**Table 1**), suggesting that statistical power was unlikely to cause this discrepancy. In the following, we focused on the CER region for a detailed comparison of AD and PSP, as this region had commonly perturbed metabolites between these two diseases, whereas TCX had none.

### 2.2 Comparison of AD and PSP metabolic signatures in CER

To compare the metabolic signatures of the two diseases in CER, each metabolite was categorized using a ‘sub-pathway’ annotation. There were 153 metabolites significantly associated with AD, 136 of these 153 were annotated with 53 pathways, and the remaining 17 metabolites were uncharacterized and thus were excluded from further analysis. Similarly, there were 35 metabolites significantly associated with PSP, 30 of which were annotated with 19 pathways, and the remaining 5 metabolites were uncharacterized and thus were excluded from further analysis. Overall, 35 pathways were exclusively dysregulated in AD, one pathway was exclusive to PSP, and 18 pathways were shared across the two diseases. These pathways were distributed across all eight super-pathways, with amino acids and lipids representing the groups with the most changes (**Figure 3**).

**Figure 3:**
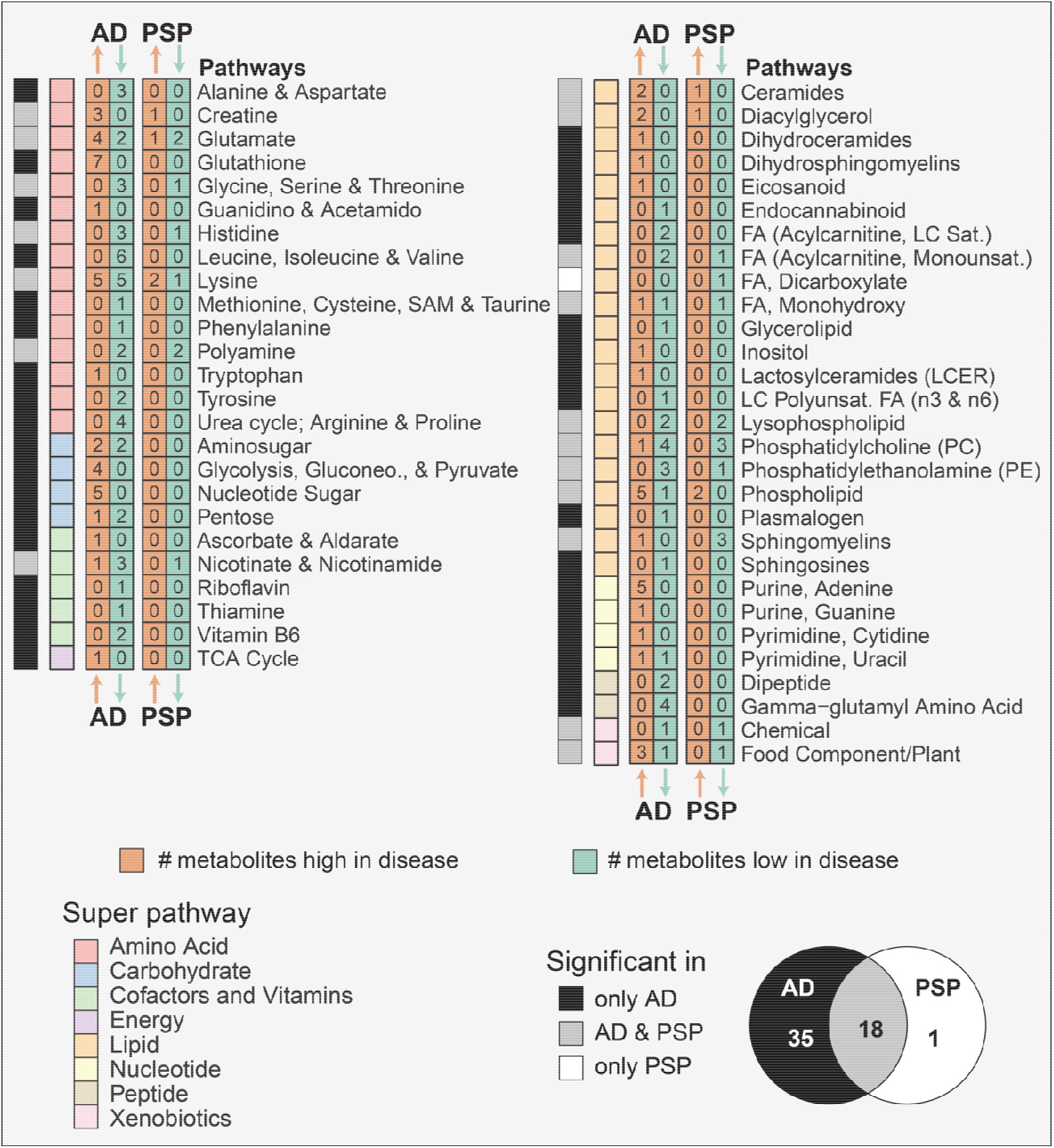
Functional annotation of metabolic signatures in CER region. Within the cerebellar cortex region, for each disease and pathway, the number of positively or negatively associated metabolites is shown in orange an green boxes respectively. These pathways are further annotated by super-pathways. The significance of these pathways in one or both diseases is shown per pathway. Venn diagram depicts the overlap of the number of significant pathways in the two diseases. FA – Fatty acid; LC – Long chain.

#### Pathways exclusively dysregulated in AD

35 pathways were dysregulated in AD but not in PSP. Among these, various amino acid pathways were negatively associated with AD including the urea cycle, alanine & aspartate, tyrosine, and branched-chain amino acid (BCAA) metabolism, while glutathione metabolism was positively associated. Further, the metabolism of vitamins B1, B2, and B6 was negatively associated with AD, while vitamin C metabolism was positively associated. Moreover, several pathways belonging to the metabolic groups of peptides, carbohydrates, energy metabolites, and nucleotide metabolism were dysregulated only in AD.

#### Pathways exclusively dysregulated in PSP

Dicarboxylate metabolism was the only pathway uniquely dysregulated in PSP. However, within this pathway, only one of the seven metabolites, the fatty acid 3-hydroxyadipate, was significantly lower in PSP as compared to controls, while changes in the remaining six metabolites were insignificant. Thus, there is limited evidence to support the dysregulation of this pathway in PSP.

#### Pathways commonly dysregulated in both diseases

18 pathways were commonly perturbed in AD and PSP. These pathways were mainly from the amino acid and lipid groups. Within the amino acid group, creatine, glutamate, and lysine metabolism were positively associated with both diseases, while polyamine, glycine, and histidine metabolism were negatively associated. Within the lipid group, ceramides, diacylglycerol, and phospholipids were positively associated with both diseases, while sphingomyelin, lysophospholipid, phosphotidylcholine, phosphoethanolamine, and FA (Acyl Carnitine, Monounsaturated) were negatively associated.

Overall, in the two diseases, common metabolic perturbations were better captured at the functional level of pathways (18/54 changed, 33.34%) than at the individual metabolite level (27/161 changed, 16.77%).

### 2.3 Functional insights into unique and shared metabolic dysregulation in AD and PSP in CER

As outlined above, in the CER region, 35 metabolic pathways were exclusively dysregulated in AD, only one pathway was exclusively dysregulated in PSP, and 18 pathways were shared across the diseases (**Figure 4**). In the following, we discuss functional insights into the commonalities and differences of metabolic dysregulation in the two diseases based on selected pathways.

**Figure 4:**
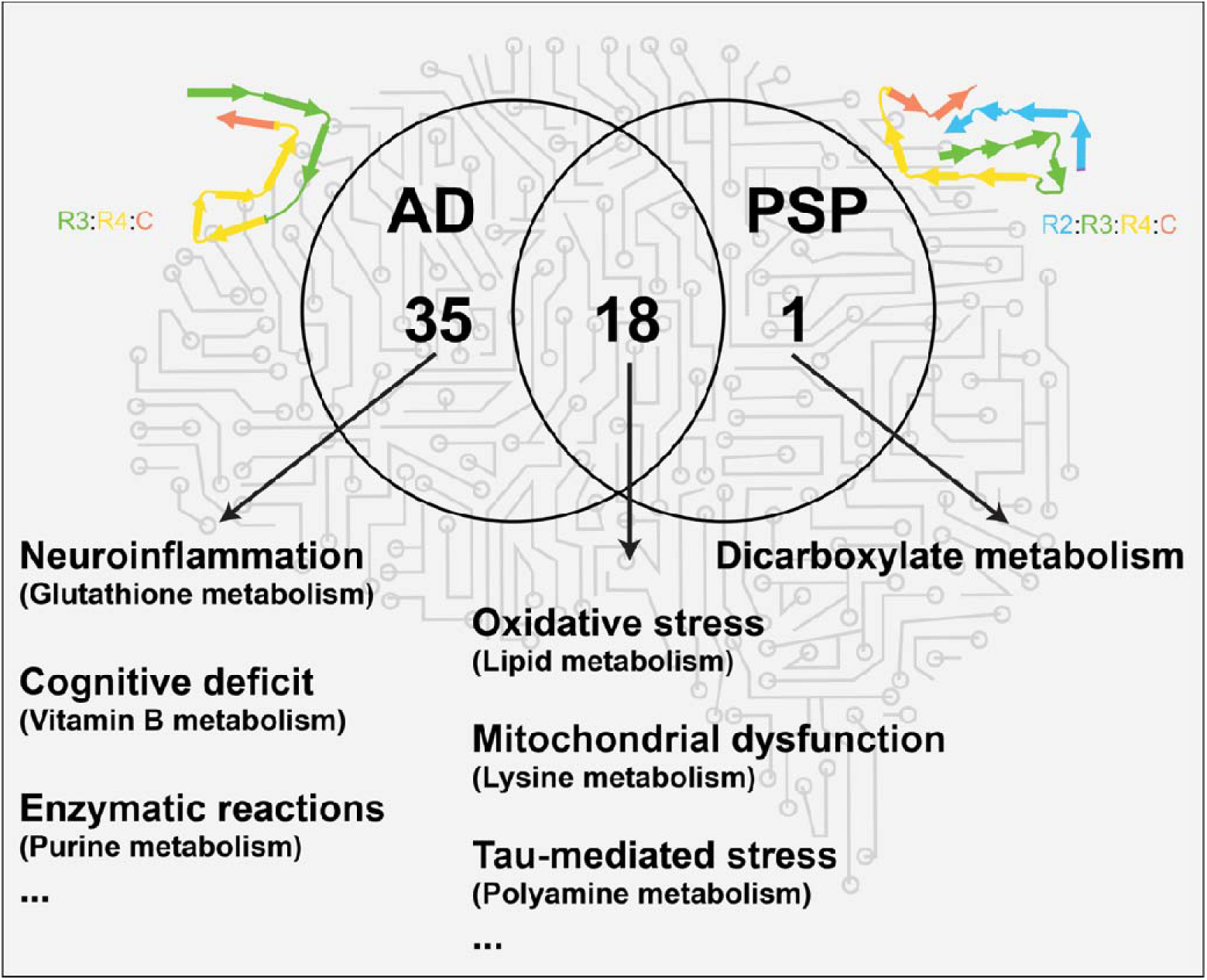
Comparison of deregulated pathways in the two diseases. Within the cerebellar cortex region, 3 pathways were exclusively dysregulated in AD, 18 pathways were shared between the two diseases, and one pathway was exclusively dysregulated in PSP. The labeled processes indicate selected pathways discussed in the text. Included for illustrative purposes are stylistic representations of the tau isoform associated with each tauopathy. Within these protein structures, residues in R1–R4 and in the C-terminal domain are colored purple, blue, green, gold, and orange, respectively^24^.

#### 2.3.1 Metabolic pathways altered exclusively in AD in CER

Within the CER region, 35 metabolic pathways were dysregulated exclusively in AD. Based on the number of metabolites altered, we highlight the three most perturbed pathways from three different metabolic groups in the following.

##### Glutathione metabolism and neuroinflammation

Neuroinflammation is central to AD pathogenesis and is marked by the release of pro-inflammatory molecules as well as reactive oxygen species^29^. Antioxidants such as glutathione play a key role in combating oxidative stress resulting from neuroinflammation^30^. Neuroinflammation and oxidative stress in the brain are believed to mediate neuronal dysfunction and death^29^. In our data, 7 out of 13 metabolites from glutathione metabolism were higher in AD as compared to controls. Of these seven, glutathione oxidized (GSSG), ophthalmate, and 4-hydroxy-nonenal-glutathione, are known biomarkers of oxidative stress^31–33^. Of note, based on genomic and transcriptomic data, we have previously demonstrated that this pathway is disrupted in AD^34^.

##### Purine metabolism and enzymatic reactions

Purine bases serve as cofactors for various biochemical reactions and are thus crucial for many metabolic processes^30^. Moreover, adenosine has been implicated in the modulation of cognition and memory, as well as the permeability of the blood-brain barrier^35^. In our data, 5 metabolites from adenine metabolism and one from guanine metabolism were higher in AD as compared to controls. Purine metabolism has been shown to be perturbed in AD; however, effect directions were inconsistent between cerebrospinal fluid and brain regions^13, 35, 36^. Overall, our findings indicate a dysregulation of purines in AD brains which has the potential to alter various brain functions.

##### Vitamin B metabolism and cognitive deficit

Multiple B vitamins are considered protective against cognitive decline^37^. In our data, thiamine (vitamin B1), riboflavin (vitamin B2), and pyridoxine (vitamin B6) were found to be lower in AD as compared to controls. Thiamine-dependent enzymes play central roles in glucose metabolism^38^. In AD brains, a reduction in thiamine levels has been associated with the diminished activity of thiamine-dependent enzymes, including transketolase from the pentose phosphate pathway and 2-ketoglutarate dehydrogenase complex from the tricarboxylic acid (TCA) cycle^38, 39^. Riboflavin and pyridoxine in their co-enzymatic forms are involved in one-carbon metabolism^40^. Riboflavin acts as a co-factor in the folate cycle, while pyridoxine is involved in the transsulphuration pathway^40^. Overall, these vitamins are essential for a well-functioning metabolism, and reduction in their levels has been associated with a decline in cognitive functions^37, 41^.

Of note, all of the above-mentioned AD-related metabolic dysregulations identified in the CER region were also detected in our previous findings in the dorsolateral prefrontal cortex region of the ROS/MAP cohort^15^.

#### 2.3.2 Metabolic pathways altered in both diseases

Within the CER region, 18 metabolic pathways were dysregulated in both diseases. In the following, we highlight pathways from the two biggest metabolic classes, lipids and amino acids.

##### Lipid metabolism and oxidative stress

In neurodegenerative diseases, lipid metabolism has been widely implicated in disease pathogenesis, including β-amyloid-associated lipid rafts, increased lipid peroxidation^42^, altered cholesterol and lipid homeostasis^43^, as well as phospholipase 2 mediated lipid cleavage^44^. In our data, lipids from various lipid classes were dysregulated, with 15 lipids dysregulated in AD and 8 of those 15 also dysregulated in PSP.

Oxidative stress, a hallmark of neurodegenerative diseases, induces lipid peroxidation, which leads to increased phospholipase activity^45, 46^. The breakdown of phospholipids by phospholipases leads to compromised membrane integrity^47^, which affects neuronal excitability^45^. In our data, several phosphatidylcholines (PCs), the major membrane phospholipids, were lower in both diseases compared to controls. Phospholipases degrade PCs into lyso-PCs, which get further broken down into glycerophoshorylcholine (GPC) and fatty acids^48^. GPC was high in both diseases as compared to controls in our study.

Moreover, oxidative stress is also known to increase ceramide production, which in turn may trigger synaptic dysfunction and neuronal death^49^. Ceramides play an important role in many cellular processes, including lipid signaling, metabolic homeostasis, and mitochondria-linked apoptosis^50^. In our data, ceramide levels were positively associated with both diseases with varying degrees. N-stearoyl-sphingadienine (d18:2/18:0)* was high in both diseases compared to controls, while lactosyl-N-stearoyl-sphingosine (d18:1/18:0)*, a lactosylceramide, and N-stearoyl-sphinganine (d18:0/18:0)*, a dihydroceramide, were high only in AD compared to controls. Moreover, the myelin sheath which assists in signal transduction across neurons is composed of ceramides^50^. Notably, in our previous transcriptome-based studies, we observed myelination dysregulation in both AD and PSP^9, 51^.

##### Lysine metabolism and mitochondrial dysfunction

Lysine is an essential amino acid that can be catabolized by two pathways: the saccharopine pathway in mitochondria and the pipecolate pathway in cytosol^52^. In our data, 10 metabolites from lysine metabolism were dysregulated. Of these, all 10 were perturbed in AD, while only 3 metabolites were altered in PSP. One of the ten metabolites was saccharopine, which was high in both AD and PSP as compared to controls. Previous studies in *C.elegans* and mice have argued that saccharopine accumulation leads to mitochondrial dysfunction^53^. Our results are thus another potential piece of evidence in support of mitochondrial dysregulation in the pathogenesis of tauopathies^54^.

##### Polyamine metabolism and tau-mediated stress

In our study, two out of eight metabolites from polyamines metabolism, spermidine and spermine, were lower in both tauopathies as compared to controls. Polyamines are known for their neuroprotective effects, such as the clearance of apoptotic cells via efferocytosis^55^ and autophagy^56^, both mechanisms are affected in AD and other neurodegenerative diseases^57^. Moreover, mouse-based studies have speculated a complex interaction between polyamines and tau neuropathology^58, 59^. Specifically, it has been hypothesized that spermidine and spermine reduce tau fibrillization, oligomerization, and tau seeding/propagation, while tau can lead to polyamine stress response^58^. The polyamine stress response is defined as the alteration in the brain polyamine metabolism that is stimulated by a stressor leading to an increase in putrescine level, while levels of spermine and spermidine decrease or remain unchanged^60, 61^. Notably, in our data putrescine levels were unchanged. Previous metabolomic studies have reported that metabolite ratios associate better with outcomes than individual metabolites^62^. Therefore, we performed a ratio analysis for the metabolites in the polyamine metabolite pathway (**Supplementary Tables 6-7**). In our analysis, ratios of putrescine/spermidine and putrescine/spermine were higher in both tauopathies as compared to controls indicating relatively higher levels of putrescine compared to the other polyamines.

## 3 Discussion

The two tauopathies AD and PSP exhibit several shared and unique characteristics. They differ in the genetic risk factors involved, tau-isoforms, main symptoms, and type of proteinopathy, with PSP being a pure tauopathy and AD exhibiting deposition of β-amyloid along with tau^1^. At the molecular level, the two diseases display similarities, particularly in the alterations of biological processes including myelination^9^ and mitochondrial metabolism^10^. However, the comparative analysis of alterations in bioenergetic homeostasis, a key feature of neurodegeneration^21^, between AD and PSP was an aspect not previously explored until this study. We provide a comparative account of metabolic alterations in AD and PSP. In a sizable cohort of 342 samples of AD, PSP or control donors from the Mayo Clinic Brain Bank, we profiled 658 metabolites in CER and TCX brain regions. 200 (30.4%) of these metabolites were significantly associated with at least one of the diseases in at least one of the regions. In the following, we discuss the key findings of our study.

Our results show that AD-related metabolic alterations encompass both regions, i.e., CER and TCX. We identified 153 AD-associated metabolic alterations within the CER region and 63 alterations within the TCX region. Remarkably, we found a larger number of metabolic alterations in CER compared to TCX. This was unexpected as CER is relatively devoid of significant gross neuropathology in AD^23^, although some pathology has been observed in the late phases of the disease^25^. The metabolic alterations observed in CER may be due to the presence of either neuropathology in late-phase AD or toxic β-amyloid/tau oligomer species. Alternatively, the metabolic changes seen in CER could reflect the modulation of CER because of the pathological changes in other regions of the brain, for example, due to altered neural network connectivity^63^. The greater number of metabolic alterations in the CER compared to the TCX could indicate the varying metabolic needs of these regions and how these needs shift with age or during neurodegeneration^64^. Further, our results indicate that PSP-related metabolic alterations were scarce in TCX (2 metabolites), while some metabolic alterations were observed in CER (35 metabolites). Since TCX has limited neuropathology in PSP^24^, our results were in line with what may be expected from this relatively unaffected region. While the CER generally shows minimal significant neuropathology in PSP^22^, it may still be susceptible to metabolic alterations due to the above-mentioned reasons.

Within the CER region, AD and PSP significantly differed in the number of metabolic alterations (153 in AD and 35 in PSP). As opposed to our previous speculation that tau is the potential driver of AD-associated metabolic alterations in the dorsolateral prefrontal cortex^15^, the present results stipulate that tau may not be the sole driver of metabolic alterations in the cerebellar cortex. Based on these results, it can be inferred that various factors differentiating these diseases may have contributed to the differences in their metabolic imprint. Furthermore, although our analysis adjusted for age as a covariate, the observed differences in age could potentially contribute to the variance in the number of metabolic alterations identified within AD and PSP brains. The median age at death of PSP donors was lower than AD donors, and controls were older than both AD and PSP cases.

Despite the differences in the two tauopathies, common metabolic processes were altered across the two diseases. Pathway analysis of the altered metabolites in the CER region indicated a higher overlap in affected biological processes (33.34%) across the pathologies as compared to individual metabolites (16.77%). We found metabolic alterations shared between the two tauopathies in various pathways and highlighted the specifics of several selected pathways. We speculate that these similarities in the two diseases can result from either common metabolic rewiring to combat the neurodegenerative processes or common perturbation of the metabolism due to the neurodegenerative process. AD showed more unique alterations, while PSP shared most of its altered processes with AD. We found that lipid metabolism-mediated oxidative stress, lysine metabolism-associated mitochondrial dysfunction, and tau-mediated polyamine stress were shared processes between the two diseases. Meanwhile, glutathione metabolism-mediated oxidative stress and neuroinflammation, purines-linked enzymatic deregulations, and vitamin B-associated cognitive deficits were unique to AD.

Our study had several limitations. First, biological and technical variations due to the postmortem interval, i.e., the time interval between death and preservation of the brain tissue, cannot be avoided in studies on postmortem brain tissue. One way to partially account for this effect is by using this interval as a confounder in the statistical models^15^. However, this was not possible in the present study due to missing data on postmortem interval. Thus, the potential degradation of certain metabolites until sample preservation could not be adjusted for. Second, bulk tissue profiling as used in this study lacks resolution at the single-cell level that might be necessary to distinguish the contribution of various cell fractions. Differential proportions of cell types or differences in microenvironment could affect bulk levels of metabolites in various ways: (1) Significant differences in cell proportion could alter the levels of metabolites in the bulk sample^65^. (2) Neuropathology could reprogram certain cell types to produce different levels of metabolites. For instance, tau accumulation within neurons could reprogram their metabolism leading to aberrations at both the metabolome and transcriptome level^9^. (3) The cellular microenvironment could influence the metabolic profiles of certain cell types. For example, pathology leads to the activation of microglial cells^67^, which could have a different metabolic profile than a microglial cell in a steady state. In summary, one or more of these mechanisms may drive the changes in metabolic profiles at the cell type-specific level. Future studies need to focus on single-cell or spatial metabolic profiling approaches to delineate these mechanisms.

In summary, we presented the first large-scale metabolic comparison of two tauopathies - AD and PSP - focusing on two different brain regions. Our study is a step toward cataloging the common and divergent metabolic changes observed in these two diseases. Follow-up studies are needed to investigate the specific metabolic pathways commonly implicated in these diseases. Moreover, the contribution of specific cell types toward the observed metabolic changes remains to be determined. Future studies with specific cell populations, using either spatial metabolomics or iPSC-derived cell type populations, will enable a precise comparison of the metabolic changes in the two diseases at the cell type specific level.

## 4 Methods

### 4.1 Cohorts, clinical data, and neuropathological data

The Mayo Clinic cohort consisted of 162 temporal cortex (TCX) and 182 cerebellar cortex (CER) samples. Among those samples, 138 TCX and CER samples were from the same brain donor. All samples received diagnoses at autopsy following neuropathologic evaluation. Briefly, sample donors with Braak neurofibrillary tangles (NFT) stage of IV or greater received a neuropathologic diagnosis of Alzheimer’s disease (AD)^23^; progressive supranuclear palsy (PSP) cases were diagnosed according to NINDS neuropathologic criteria^68^; control samples were defined by a Braak NFT stage of III or less, CERAD neuritic and cortical plaque densities of 0 (none) or 1 (sparse), and lack of any of the following pathologic diagnoses: AD, Parkinson’s disease (PD), dementia with Lewy bodies (DLB), vascular dementia (VaD), PSP, motor neuron disease (MND), corticobasal degeneration (CBD), Pick’s disease (PiD), Huntington’s disease (HD), FTLD, hippocampal sclerosis (HipScl) or dementia lacking distinctive histology (DLDH). All 378 sample donors were of North Americans of European descent. Details on this cohort have been provided in previous studies^9, 10, 69^.

### 4.2 Metabolomics profiling

The untargeted HD4 metabolomics platform from Metabolon Inc. was used to measure brain metabolic profiles. Fractions of the tissue samples were used for two ultra-high performance liquid chromatography-tandem mass spectrometry (UPLC-MS/MS; positive ionization), a UPLC-MS/MS (negative ionization), and a UPLC-MS/MS polar platform (negative ionization). Based on the spectra, area under the curve was used to quantify the peak intensities. An internal spectral database was used for compound identification. Further details can be found in a previous publication^15^.

### 4.3 Data preprocessing

A total of 827 metabolites were identified by the metabolomics platform. Of these, 658 metabolites with <25% missing values were used for the analysis, the remaining 169 were excluded. To correct for sample-wise variation, probabilistic quotient normalization was performed^70^, followed by log_2_ transformation. To impute the remaining missing values, a k-nearest-neighbor-based algorithm was applied as described previously^71^. The local outlier factor method was used for outlier detection^72^, identifying no samples to be excluded. 278 irregularly high or low single concentrations with absolute z-score above *q = abs(qnorm(0.0125/n*)), with *n* representing the number of samples, were set to missing. This formula finds the cutoff for values with less than 2.5% two-tailed probability to originate from the same normal distribution as the rest of the measurement values, after applying a Bonferroni-inspired correction factor (division by sample size). These values were then imputed using a k-nearest-neighbor-based algorithm^15^.

### 4.4 Differential analysis of metabolites

342 out of 344 samples were used for the analysis; 2 with missing *APOE* ε4 status were excluded. To identify metabolites associated with diagnosis, two subsequent logistic regressions were used as described previously^73^. The first model included diagnosis as the outcome, metabolites as the predictors, but no confounding factors. P-values of this model were corrected using the Benjamini-Hochberg (BH) method^74^ for multiple hypothesis testing. The second logistic regression model included diagnosis as the outcome, metabolites with adjusted p-values < 0.25 from the first model as predictors, and sex, number of *APOE* ε4 alleles, and age at death as confounders. Metabolites with p-value < 0.05 in the second model were used to define the metabolic signatures of the diseases. All analyses were performed using the maplet R package^75^.

For a selected set of metabolites i.e., from polyamine metabolism, pairwise ratios were computed (Supplementary Tables 6-7). To this end, exponentials of the log_2_ transformed metabolic profiles were computed, ratios were generated, and then log_2_ transformed. The differential analysis of these ratios was performed in the same way as individual metabolites.

## Ethics approval and consent to participate

This study was approved by the appropriate Mayo Clinic Institutional Review Board. All participants or next-of-kin provided consent.

## Data availability

Metabolomics as well as clinical data for the Mayo Clinic cohorts are available via the AD Knowledge Portal. The AD Knowledge Portal is a platform for accessing data, analyses, and tools generated by the Accelerating Medicines Partnership (AMP-AD) Target Discovery Program and other National Institute on Aging (NIA)-supported programs to enable open-science practices and accelerate translational learning. The data, analyses, and tools are shared early in the research cycle without a publication embargo on a secondary use. Data is available for general research use according to the following requirements for data access and data attribution (https://adknowledgeportal.org/DataAccess/Instructions). For access to the metabolomics data used in this manuscript see: https://www.synapse.org/#!Synapse:syn26446587

An interactive view of AD associations from this study can be found at https://krumsieklab.shinyapps.io/tauopathies/

All R scripts to generate the tables and figures of this paper are available at https://github.com/krumsieklab/ad-mayo-tauopathies

## Acknowledgments

We thank the patients and families for their participation, without whom these studies would not have been possible.

This work was done as part of the National Institute of Aging’s Accelerating Medicines Partnership for AD (AMP-AD) and was supported by NIH grants 1U19AG063744, 1R01AG069901-01A1, U01AG061357, P30AG10161, P30AG72975, R01AG15819, R01AG17917, U01AG46152, U01AG61356, RF1AG058942, RF1AG059093, and U01AG061359. The results published here are in whole or in part based on data obtained from the AD Knowledge Portal (https://adknowledgeportal.org).

The Mayo Clinic samples are part of the RNAseq study data led by Dr. Nilüfer Ertekin-Taner, Mayo Clinic, Jacksonville, FL as part of the multi-PI U01 AG046139 (MPIs Golde, Ertekin-Taner, Younkin, Price). Samples were provided from the following sources: The Mayo Clinic Brain Bank. Data collection was supported through funding by NIA grants P50 AG016574, R01 AG032990, U01 AG046139, R01 AG018023, U01 AG006576, U01 AG006786, R01 AG025711, R01 AG017216, R01 AG003949, NINDS grant R01 NS080820, CurePSP Foundation, and support from Mayo Foundation. Study data includes samples collected through the Sun Health Research Institute Brain and Body Donation Program of Sun City, Arizona. The Brain and Body Donation Program is supported by the National Institute of Neurological Disorders and Stroke (U24 NS072026 National Brain and Tissue Resource for Parkinson’s Disease and Related Disorders), the National Institute on Aging (P30 AG19610 Arizona Alzheimers Disease Core Center), the Arizona Department of Health Services (contract 211002, Arizona Alzheimers Research Center), the Arizona Biomedical Research Commission (contracts 4001, 0011, 05-901 and 1001 to the Arizona Parkinson’s Disease Consortium) and the Michael J. Fox Foundation for Parkinson’s Research.

RB is also supported by Alzheimer’s association award AARFD-22-974775. RB thanks her colleagues from the Krumsiek lab for fruitful discussions and support in this work. NET, XW and MA are supported by R01AG061796, U01AG046139 and U19AG074879. NET is also supported by the Alzheimer’s Association Zenith Award.

## Author contributions

RB, JK, XW, NET, and RKD designed the study and analytic approaches. RB and JK designed the computational and statistical methods, performed the analysis, interpreted the results, and drafted the manuscript. MAllen, MArnold, and GK contributed to scientific discussions. NET, XW, MAllen provided samples and phenotypic data on the Mayo Clinic cohort, interpreted the results, edited the manuscript. CB curated and managed data. NET acquired funding, supervision and overall direction for the Mayo Clinic cohort. RKD acquired funding and is the overall PI of the Alzheimer’s disease metabolomics consortium. All authors read and reviewed the manuscript.

